# Mucosal-associated invariant T (MAIT) cell responses in *Salmonella enterica* serovar Typhi strain Ty21a oral vaccine recipients

**DOI:** 10.1101/2022.10.04.22280651

**Authors:** Shubhanshi Trivedi, Olivia Cheng, Ben J Brintz, Richelle C. Charles, Daniel T Leung

## Abstract

**Background:** Mucosal–associated invariant T (MAIT) cells are unconventional innate-like T cells abundant in mucosal tissue of humans, and associated with protective responses to microbial infections. MAIT cells have capacity for rapid effector function, including the secretion of cytokines and cytotoxic molecules. However, limited information is available regarding the activity of MAIT cells in mucosal vaccine-mediated immune responses in humans.

**Methods:** We enrolled healthy adults who received a course of oral live-attenuated *S*. Typhi strain Ty21a vaccine and collected peripheral blood samples pre-vaccination, and at 7 days and one month post-vaccination. We used flow cytometry, cell migration assays, and tetramer decay assay to assess MAIT cell responses.

**Results:** We show that following vaccination, circulating MAIT cells are decreased in frequency but have increased activation markers. Post-vaccine timepoints had higher levels of MAIT cells expressing gut-homing marker integrin α4β7 and chemokine receptor CCR9, suggesting the potential of MAIT cells to migrate to mucosal sites. While we found higher frequencies of TNF-α expression on MAIT cells post-vaccination, we did not find significant differences in expression of other effector molecules, TCR avidity, or cell migration.

**Conclusions:** We show how MAIT cell immune responses are modulated post-vaccination against *S*.Typhi. This study contributes to our understanding of MAIT cells’ potential role in oral vaccination against bacterial mucosal pathogens.

## Introduction

Mucosal-associated invariant T (MAIT) cells are innate-like αβ T cells defined by the expression of an invariant α chain, generally Vα7.2 linked to Jα33, Jα12, or Jα20 in humans and a limited array of T cell receptor β (TCRβ) chains [1, 2]. MAIT cells are restricted by the non-classical MHC-related molecule 1 (MR1) and respond to vitamin B metabolites derived from bacterial and fungal species [3]. MAIT cells have been associated with protection and antibacterial immune defense in various bacterial infections, including *Legionella longbeachae, Mycobacterium tuberculosis* (Mtb), *Mycobacterium bovis, Francisella tularensis, Escherichia coli, Vibrio cholerae*, and *Klebsiella pneumoniae* [4-8]. In human challenge studies, exposure to enteric bacteria such as *Salmonella enterica* serovar Typhi (*S*. Typhi), *S*. Paratyphi A, and *Shigella flexneri* resulted in MAIT cells activation, proliferation, and homing to mucosal sites [9-11]. MAIT cells functions include their capacity to secrete TNF-α, IFN-γ, IL-17, as well as Granzyme B [12, 13]. MAIT cells are attractive vaccine targets [14] as they bridge the adaptive and innate immune responses against bacterial infections and are donor-unrestricted (not restricted by MHC polymorphism) [15].

Few studies have described MAIT cell activity during oral vaccination. In volunteers vaccinated with an attenuated strain of *Shigella dysenteriae*, MAIT cell activation was seen in those who mounted LPS-specific IgA antibody-secreting cell responses [10]. Specific to *S*. Typhi infection outside of vaccination, human challenge studies with *S*. Typhi have shown that MAIT cells are activated, exhausted and depleted during infection [11]. However, there exists a lack of knowledge regarding the activity and function of MAIT cells in humans following mucosal vaccination, such as with oral attenuated *S*. Typhi strain Ty21a, a commercially available oral mucosal vaccine against typhoid fever [16]. The objective of our study was to examine longitudinal MAIT cells responses in a cohort of Ty21a recipients. We show that Ty21a vaccination results in changes in MAIT cell frequency, activation, cytokine production and homing markers expression.

## Materials and Methods

### Subjects

Eleven healthy volunteers, six males (aged 42 ± 15 years) and 5 females (aged 40 ± 10 years) participated in this study. All volunteers provided written informed consent. Four doses of a single oral capsule of Ty21a (Vivotif, PaxVax) were taken on days 1, 3, 5 and 7. Blood samples were collected upon study enrollment (Day 1) and again in the same subject’s approximately 7 days and 1 month after the last dose of vaccine (Day 7 and 1 month post-vaccination, p.v.). Venous blood was collected and centrifuged over a Lymphoprep (STEMCELL Technologies Inc) density gradient using a standard protocol to isolate peripheral blood mononuclear cells (PBMCs). PBMCs in cryogenic vials were placed immediately into an isopropanol freezing container (Nalgene Mr. Frosty) and were cryopreserved in -80°C until use for immunologic assays. Study procedures were reviewed and approved by the Institutional Review Board of the University of Utah (IRB #84287).

### Antigenic stimulation and incubation

Viable *S*. Typhi Ty21a were obtained by dissolving a vaccine capsule (Vivotif) in 10 ml brain heart infusion (BHI) media and incubating overnight at 37°C. The bacteria were then subcultured (1:10) in BHI media for four hours (O.D = 0.4), harvested and stored in 50% glycerol at -80°C. For phenotypic analysis of MAIT cells, PBMCs were thawed from −80°C and were seeded (2 × 10^6^ cells/well) in complete medium in 96-well U bottom plates. Cells in each well were stimulated at 100 multiplicity of infection (MOI) with heat-killed *S*. Typhi Ty21a (killed by incubation at 95°C for 30 min). Unstimulated control wells were treated with complete medium. Cells were then incubated at 37° in 5% CO_2_. After an overnight incubation, 1X brefeldin A (BD GolgiPlug; BD Biosciences) was added to each well, and the plate was incubated for a further 4 hours at 37° in 5% CO_2_.

### Flow cytometry

PBMCs were washed before being stained for viability and surface phenotype. For intracellular cytokine analysis, surface staining was followed by fixation and permeabilization and staining using foxp3/transcription factor staining buffer set (eBioscience). Details of the antibodies that were used are presented in **Supplementary table 1**. Compensation beads (BD Biosciences) were used to create compensation matrices, and Fluorescence minus One (FMO) controls were used to identify populations of interest. All samples were acquired using Cytek Aurora (Cytek Biosciences) and analyzed using FlowJo software v10 (Tree Star Inc, Ashland, OR).

### Cell migration assay

To assess migratory properties of MAIT cells, PBMCs were stimulated with heat-killed *S*. Typhi Ty21a overnight, washed, and resuspended in RPMI 1640 medium supplemented with 0.1% bovine serum albumin (Thermo Fisher Scientific). Cells were seeded in the upper chamber of a 6-well transwell plate inserts with a pore size of 3 μm (Thermo Fisher Scientific) at a density of 1 × 10^6^ cells/well. Cells were allowed to migrate against a gradient of 150 ng/ml recombinant chemokines CCL20/MIP-3α and CCL25/TECK-3 (Peprotech) in RPMI 1640 medium supplemented with 0.1% bovine serum albumin for 4 hours at 37°C [17]. Cells migrated into the bottom chamber were collected, washed in FACS buffer (phosphate-buffered saline with 2% fetal bovine serum), stained with fluorochrome-conjugated anti-CD8a, anti-TCRVα7.2, anti-CD161 antibodies and anti-human MR1 5-OP-RU Tetramer (NIH Tetramer Core Facility), and counted using Countbright^™^ absolute counting beads (Thermo Fisher Scientific) and flow cytometry.

### Tetramer staining and decay assay

Tetramer staining and decay assay were performed as previously mentioned [9]. Briefly, for PBMCs, cells were stained with live/dead fixable viability dye efluor 780 (eBiosciences) for 15 minutes at room temperature (RT), followed by incubation with 5-OP-RU MR1 tetramer for 40 min at RT. Excess tetramer was washed off and cells re-suspended in FACS buffer with or without 20 μg/ml anti-MR1 antibody (clone 26.5, Biolegend). Samples were left at 37°C and periodic samples were taken, washed and fixed immediately. CD3 staining was performed after collecting all samples and analyzed using flow-cytometry.

### ELISAs

For enzyme-linked immunosorbent assays (ELISA), we used *S*. Typhi LPS antigen and assessed plasma antibody responses (immunoglobulin [Ig] M, IgA, and IgG). ELISAs were performed as previously described [18]. Briefly, microplates (nunc-maxisorp flat-bottom 96-well plates, Invitrogen) were coated with 1 μg/ml LPS, and plasma was added at a dilution of 1:200 for IgG and 1:100 for IgA and IgM. Bound antibodies were detected with anti-human IgG, IgA, and IgM conjugated with horseradish peroxidase (Jackson ImmunoResearch), and plates were developed by adding 100 μl/well TRB substrate for 10 min in the dark. Development was stopped by the addition of 100 μl/well of 0.2N H_2_SO_4_ and OD read at 450 nm in an ELISA microplate reader (Multiskan Ascent; Thermo Labsystems).

### Statistical analysis

Paired comparisons were made using Wilcoxon matched-pairs signed-rank test using Prism v9 (GraphPad). P values are two-tailed and considered significant at P < 0.05. Given the lack of prior studies on MAIT cells in oral vaccination, we were unable to perform a power calculation to inform sample size.

## Results

### Vaccination resulted in decrease in frequency of circulating MAIT cells but an increase in markers of MAIT activation

Analysis of plasma antibody responses to *S*. Typhi LPS in Ty21a vaccine recipients revealed that vaccine elicited significantly higher IgA and IgG antibody responses at 7 days and 1 month post-vaccination compared to day 1 (pre-vaccination) (Figure 1A and B). No significant differences were found in IgM antibody responses (Figure 1C). These results are consistent with prior studies of Ty21a vaccination [19-21].

**Figure 1.**
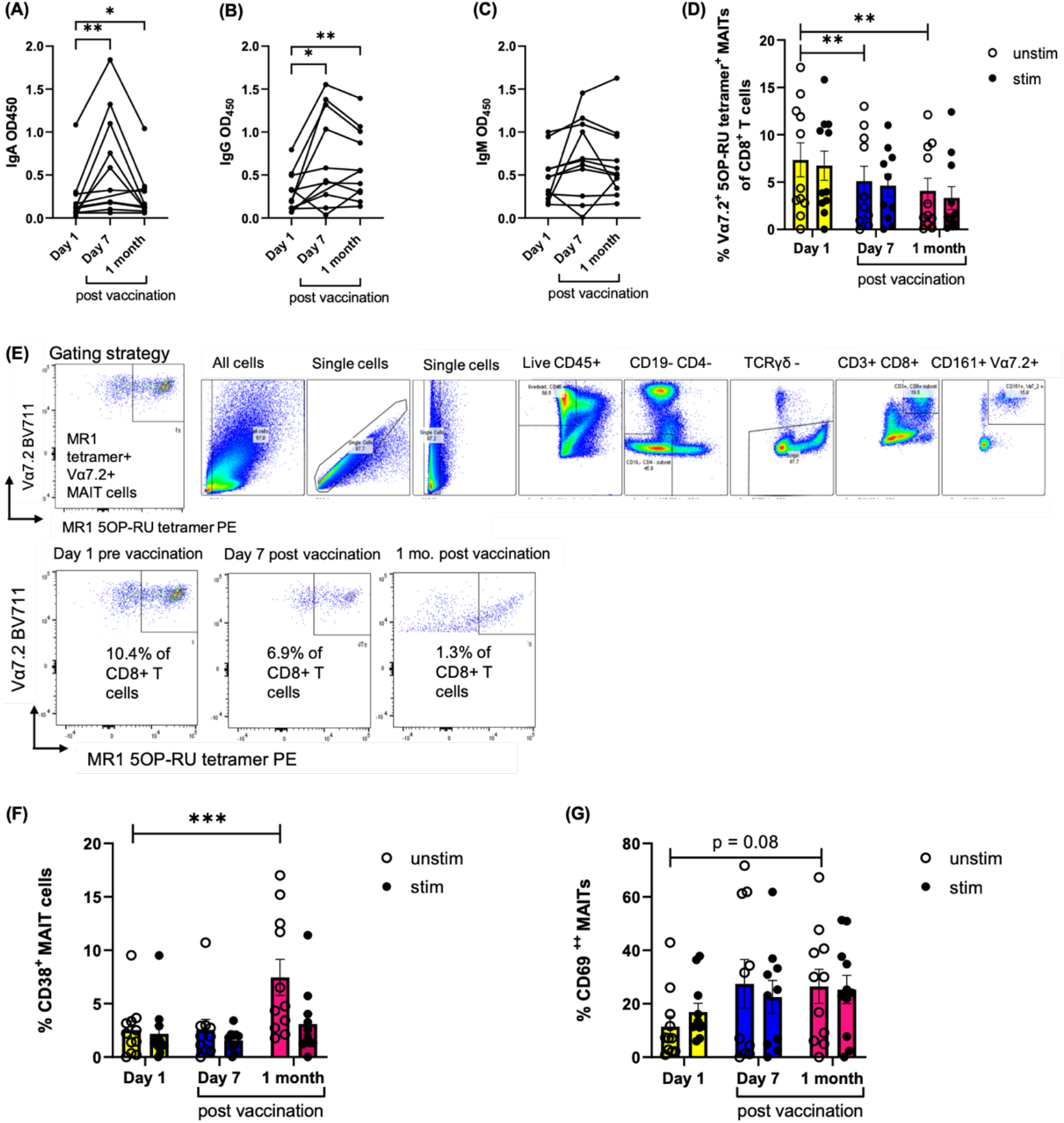
*S*. Typhi-specific antibody response, MAIT cell frequency, and MAIT cell activation after Ty21a vaccination. Plasma antibodies against *S*. Typhi LPS IgA **(A)**, IgG **(B)** and IgM **(C)** antibody responses pre-vaccination (day 1) and post-vaccination (day 7 and 1 month). **(D)** MAIT cells frequency as percentage of CD8^+^ T cells in *invitro* heat-killed *S*. Typhi Ty21a vaccine antigen-stimulated samples and unstimulated controls. **(E)** MAIT cell gating strategy and representative flow cytometry plots. **(F and G)** Frequency of activated MAIT cells expressing CD38 and CD69 pre and post-vaccination. Data were expressed as mean ± SEM of two independent experiments. \**P* < .05, \*\**P* < .01, \*\*\**P* < .001 in Wilcoxon signed-rank test (paired samples).

Because of the potential importance of MAIT cells against *S*. Typhi infections and their potential as targets for vaccine development, we determined whether immunization with the Ty21a typhoid vaccine elicits MAIT cell immune responses. PBMCs from Ty21a vaccinees were isolated and stimulated with heat-killed *S*. Typhi Ty21a, and MAIT cells were analyzed using flow cytometry. Defining MAIT cells as live CD45^+^ CD19^-^ TCRγd^-^ CD4^-^ CD3^+^ CD8^+^ CD161^+^ Vα7.2^+^ MR1 5OP-RU tetramer^+^, we found that the frequencies of circulating MAIT cells as a proportion of total CD8+ cells significantly decreased in unstimulated and stimulated conditions both at day 7 and at one month post-vaccination compared to pre-vaccination (Figure 1D and E). We next measured the expression of activation markers CD38 and CD69 in MAIT cells. We found that the frequencies of CD38^+^ MAIT cells (Figure 1F) and CD69^+^ activated MAIT cells (Figure 1G) were increased at 1 month post-vaccination. Conventional CD4+ and CD8+ T cells frequencies increased post-vaccination, consistent with a previous publication, [22] and the frequencies of CD14+ monocytes decreased in circulation post-vaccination (supplementary figure 1). No significant differences were found in B cells, CD4+ CD8+ MAIT cells, CD4+ MAIT cells and TCRγδ cells post-vaccination compared to pre vaccination (supplementary figure 1). Taken together, we found that human vaccination with Ty21a impacted the frequency and activation of circulating CD8+ MAIT cells.

### Vaccination resulted in higher frequencies of TNFα expressing circulating MAIT cells, but not of other effector molecules

We next examined whether vaccination with Ty21a induces MAIT cells to express effector molecules such as IFN-γ, TNF-α, IL-17a and cytotoxic molecules. Using flow cytometry, we found a significant increase in the frequencies of MAIT cells expressing TNF-α post-vaccination (Fig 2A), and IFN-γ expressing MAIT cells also were non-significantly higher (p = 0.08) post-vaccination in unstimulated conditions (Fig 2B). No significant differences were found in IL-17a, granzyme B, or perforin among the groups (Figure 2 C-E). IFN-γ, TNF-α and perforin expressing MAIT cells significantly increased in response to *ex-vivo S*.Typhi stimulation compared to unstimulated condition at the pre-vaccination timepoint. However, except for TNF-α at 1 month post-vaccination, we did not observe any significant differences between unstimulated and stimulated conditions at any of the post-vaccination timepoints (Figure 2A).

**Figure 2.**
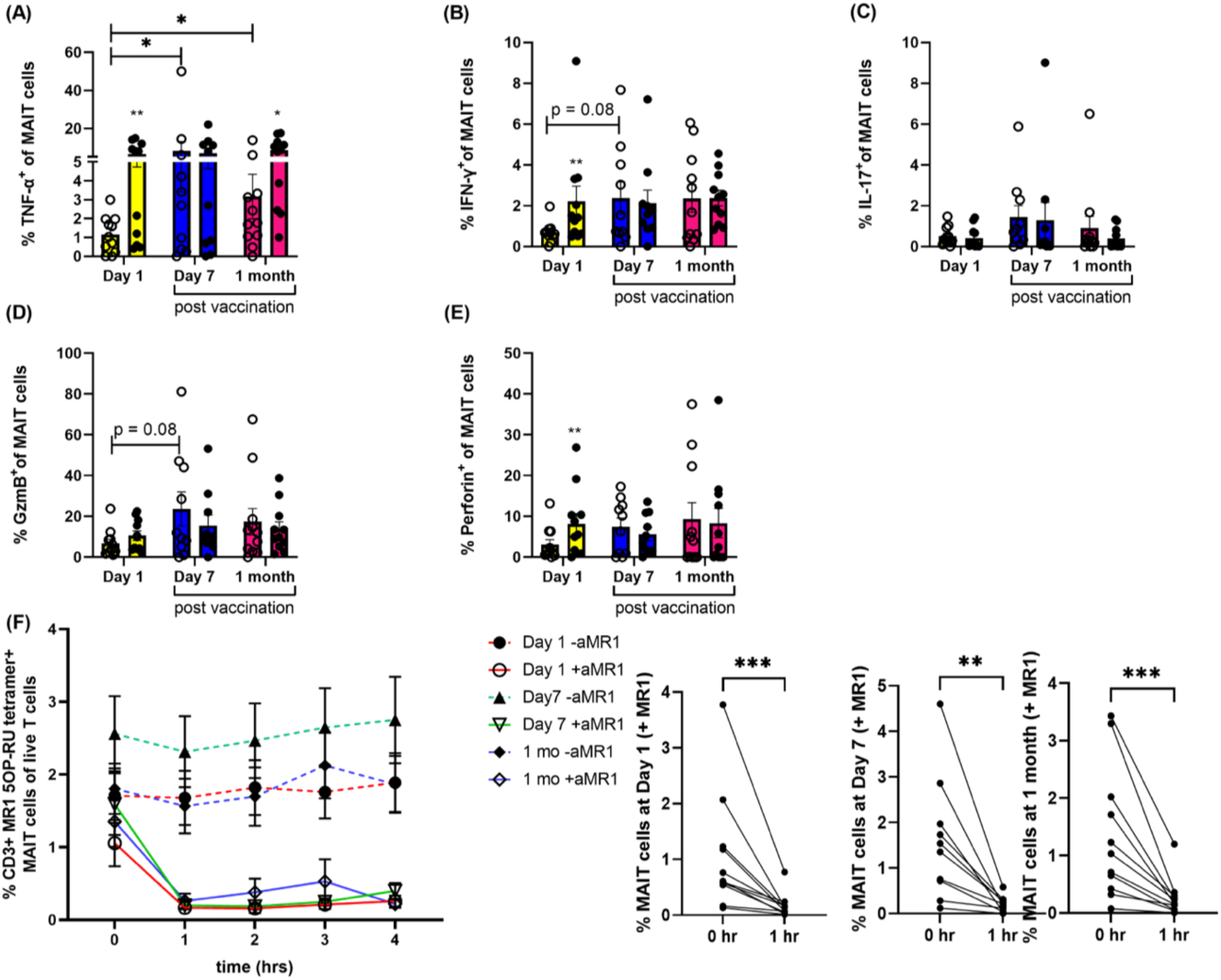
MAIT cell effector function and tetramer dissociation post vaccination. PBMCs obtained from vaccine recipients (n = 11 per group) were stimulated with *S*. Typhi at moi of 100 and intracellular expression of **(A)** TNF-α **(B)** IFN-γ, **(C)** IL-17, **(D)** granzyme B and **(E)** perforin by MAIT cells were analyzed using flow cytometry (open circle shows unstimulated and closed circle shows stimulated data). **(F)** MAIT cell tetramer decay with (solid lines) and without (dashed lines) the presence of 20 μg/mL MR1 blocking antibody (aMR1) is plotted as percentages of live CD3+ T cells pre and post vaccination. Data were expressed as mean ± SEM of two independent experiments. Paired comparisons were made using Wilcoxon matched-pairs signed-rank test using Prism v9 (GraphPad).

### Vaccination did not result in changes in circulating MAIT TCR avidity

To determine whether the MAIT cell population’s pre- and post-vaccination display different TCR avidities for their 5OP-RU–MR1 complexes, we stained PBMCs with 5OP-RU tetramers, and assayed the rate of tetramer dissociation by incubating cells over a time course with an anti–MR1 blocking antibody [9]. In the presence of anti-MR1 antibody, tetramer dissociated rapidly over an hour interval in both pre- and post-vaccination groups. The percentage of MAIT cells bound to tetramer were similar in pre-vaccinated and post-vaccinated samples, indicating no difference in MAIT TCR avidity following vaccination (Figure 2F).

### Increased tissue-homing chemokine receptors and integrin expression on MAIT cells post Ty21a vaccination

Given that we found lower frequencies of circulating MAIT cells post-vaccination, it is possible that MAIT cells home to other compartments, such as the gut or other mucosal tissues. To address this possibility, we measured the expression of integrin α4β7 and CCR9 molecules, gut-homing markers known to be found on MAIT cells [23-26]. We found that the proportion of MAIT cells expressing integrin α4β7 increases at day 7 post-vaccination upon stimulation (Fig 3A), and the proportion of MAIT cells expressing CCR9 were higher one month post-vaccination in unstimulated and stimulated conditions (Figure 3B). Next, we used a trans-well chemotaxis assay to assess the potential migration of MAIT cells in the presence of CCR9 ligand CCL25/TECK-3, and found higher MAIT cell migration CCL2 at one month post-vaccination, though this analysis was limited by sample size due to inadequacy of sufficient PBMCs at this later time point (p = 0.25, supplementary figure 2). We also found higher percentages of MAIT cells expressing CCR4, CCR5 and CCR6 chemokine receptors (involved in tissue homing [12, 27-29]) one month post-vaccination (Figure 3 C-E). No significant differences were observed in MAIT cells expressing CD103/αE β7 integrin (skin/liver homing [23]) expression (Figure 3F) and MAIT cell chemotaxis for CCR6 ligand MIP-3α (supplementary figure 2). Furthermore, given recent identification of CXCR5+ follicular helper-like MAIT cells [30], we found a higher proportion of stimulated MAIT cells at post-vaccination time points to be positive for CXCR5^+^ (Figure 3G). Taken together, we found evidence that the decreased proportion of MAIT cells in blood may be associated with increased homing and migration into tissues following Ty21a vaccination.

**Figure 3.**
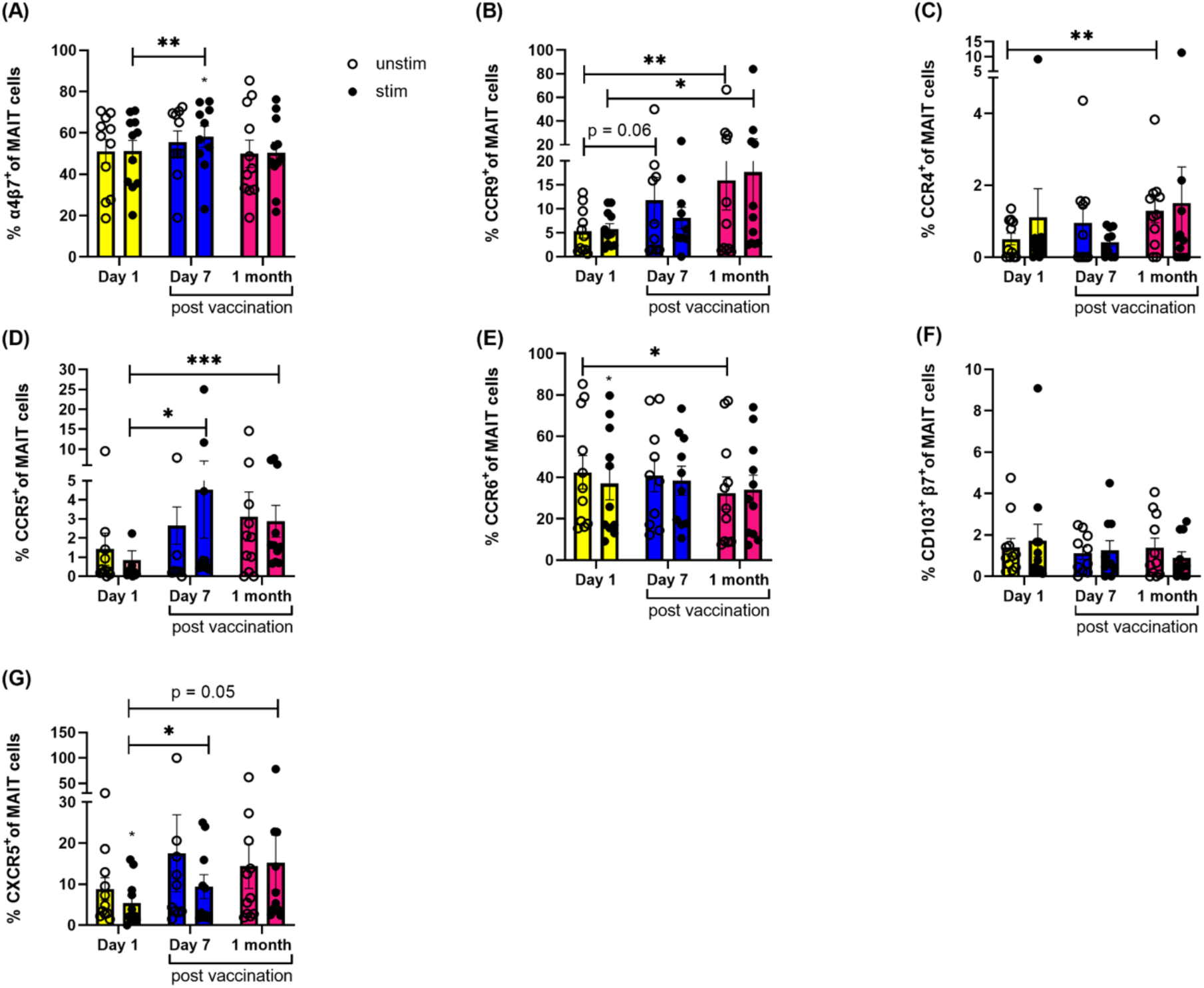
Increased homing markers and chemokine receptor expression in MAIT cells post vaccination. PBMCs obtained from vaccine recipients (n = 11 per group) were stimulated with *S*. Typhi at moi of 100 and surface expression of **(A)** Integrin α4β7, **(B)** CCR9, **(C)** CD103, **(D)** CCR4, **(E)** CCR5, **(F)** CCR6, **(G)** CXCR5 in MAIT cells was measured using flow cytometry. Data were expressed as mean ± SEM of two independent experiments. Paired comparisons were made using Wilcoxon matched-pairs signed-rank test.

## Discussion

MAIT cells have been implicated in protective responses to microbial infections and as a result could be an attractive vaccine target because they are not restricted by donor genotype, are relatively abundant in humans, and have capacity for rapid effector function [14, 15]. While studies have described MAIT cell responses to systemic vaccination in humans [10, 31, 32], mice [8] and macaques [33], there are very limited data on how MAIT cells respond to mucosal vaccines. In this longitudinal study, we evaluated MAIT cell response 7 days and one month after *S*. Typhi strain Ty21a vaccination, and found changes in circulating MAIT cell frequency, activation and homing markers.

We found that circulating MAIT cells are reduced in frequency and activated following Ty21a vaccination. Previous studies have shown that in humans, MAIT cells are lower in frequencies in the blood of children following *Vibrio cholerae* infection [7], patients with active *Mycobacterium tuberculosis* (TB) [34], and people living with human immunodeficiency virus (HIV) infection [35]. Our results are in line with a prior study demonstrating that oral challenge of volunteers with wild-type *S*. Typhi results in a sharp decline of circulating MAIT cells 48 and 96 h after typhoid diagnosis, with MAIT cells highly activated and co-expressing CCR6 and CCR9 homing markers [11]. Our results are also consistent with a prior study of subjects receiving an attenuated *Shigella* vaccine [10]. One possible explanation for lower frequencies of MAIT cells in circulation is that they migrate to mucosal tissues, where they may be recruited to contribute to innate effector responses [36]. In support of this, we found that MAIT cells post-vaccination had increased expression of several mucosal-homing chemokine receptors, including CCR6, CCR9 and integrin α4β7. Further work is needed to determine the contribution and activity of MAIT cells localized to the gut post-Ty21a vaccination.

MAIT cells produce effector molecules such as IFN-γ, TNF-α and cytotoxic molecules required for killing and eliminating bacteria-infected cells. In our study, we found increased frequencies of TNF-α expressing circulating MAIT cells post-vaccination but no significant differences in cytotoxic molecules. However, tissue-resident MAIT cells can have different phenotype and cytokine production than those in the circulation [37, 38]. We also found that while pre-vaccination samples showed significant increases in IFN-γ and perforin, upon ex-vivo stimulation with heat-killed *S*. Typhi, this was not seen in post-vaccination samples. This could be because MAIT cells in vaccinees might have been activated with Ty21a vaccine *in-vivo* (“primed”) and this baseline may not get enhanced after stimulation because of less availability of unbound MAIT-TCR. This observed “priming” of MAIT cells to mount increased expression of cytokines and cytotoxic molecules in the unstimulated samples has also been detected in pregnancy [39].

Our study has a number of limitations. First, due to the preliminary nature of this study, we had a small sample size, though this did not preclude us in observing significant changes in MAIT cell response post-Ty21a vaccination. Studies with larger cohorts are required to confirm and expand our findings. Secondly, our study was limited to description of MAIT cells in blood; given our findings of increased homing markers and migration, future studies examining tissue-resident MAIT cells are needed. Despite these shortcomings, data from this study has added to the limited body of evidence on the MAIT cell response to oral vaccination. Given the burden of disease of enteric infections, there is an urgent need of developing more effective mucosal vaccines, including orally administered ones [40]. Our findings are the first step towards harnessing MAIT cells function, which may offer an important therapeutic strategy to improve mucosal immunity.

## Data Availability

All data produced in the present work are contained in the manuscript.

## Acknowledgments

This research was supported by the National Institutes of Health (AI130378 to D.T.L.). We would like to thank the study volunteers for the blood samples. We would also like to thank the staff of the University of Utah Flow Cytometry Core.

## Supplemental Figures

**Supplementary Figure 1.**
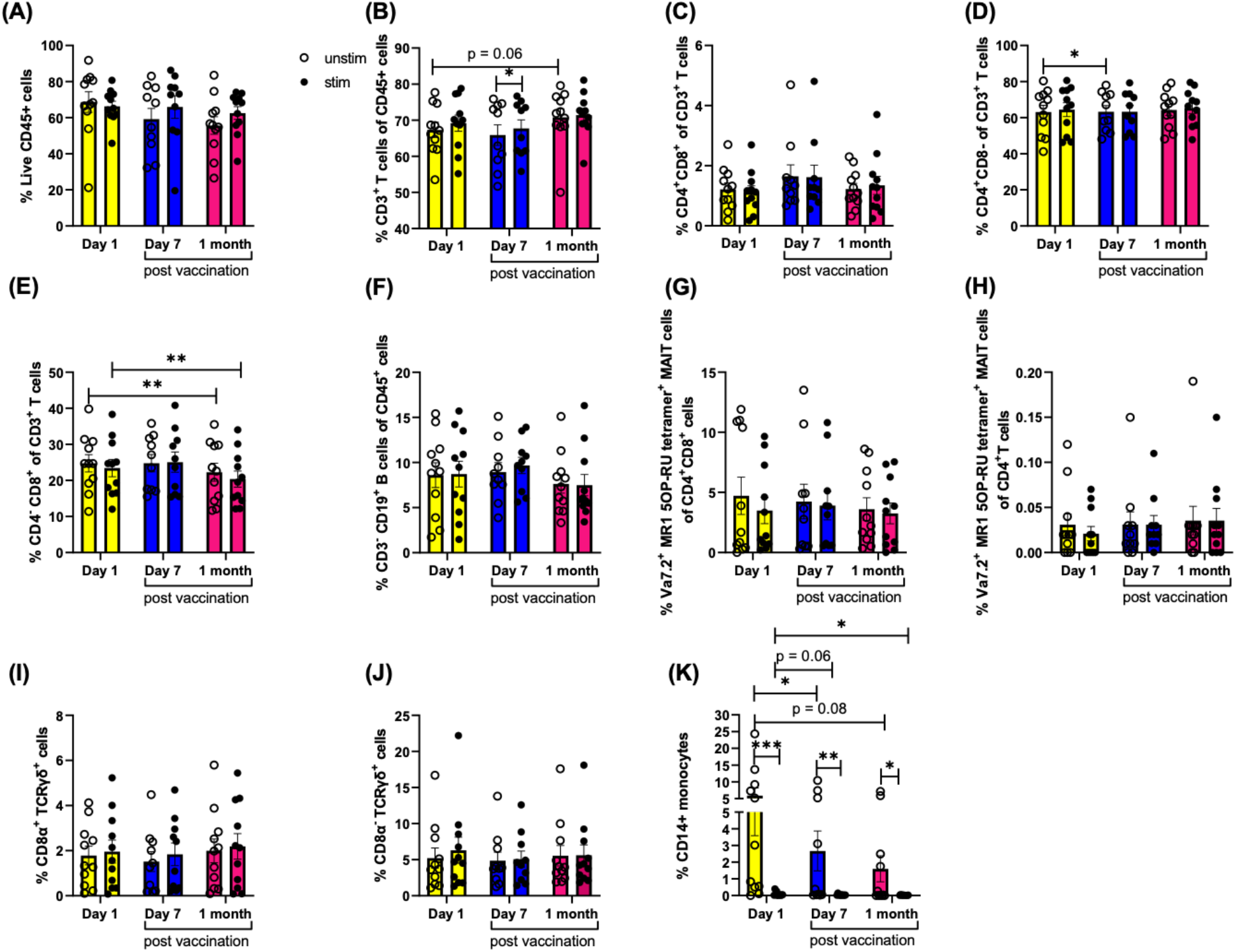
Effect of Ty21a vaccination on CD4+CD8+ MAIT cells, CD4+ MAITs and other non-MAIT populations. Frequencies of **(A)** CD45+ cells **(B)** CD3+ T cells, **(C)** CD4+ CD8+ T cells, **(D)** CD4+ T cells, **(E)** CD8+ T cells, **(F)** CD19+ B cells, **(G)** CD4+CD8+ MAIT cells, **(H)** CD4+ MAIT cells, **(I)** CD8+ TCRγδ+ cells, **(J)** CD8-TCRγδ+ cells and **(K)** CD14+ monocytes at day 1 (pre-vaccination) and day 7 and one month post vaccination. Data were expressed as mean ± SEM of two independent experiments. Paired comparisons were made using Wilcoxon matched-pairs signed-rank test.

**Supplementary Figure 2.**
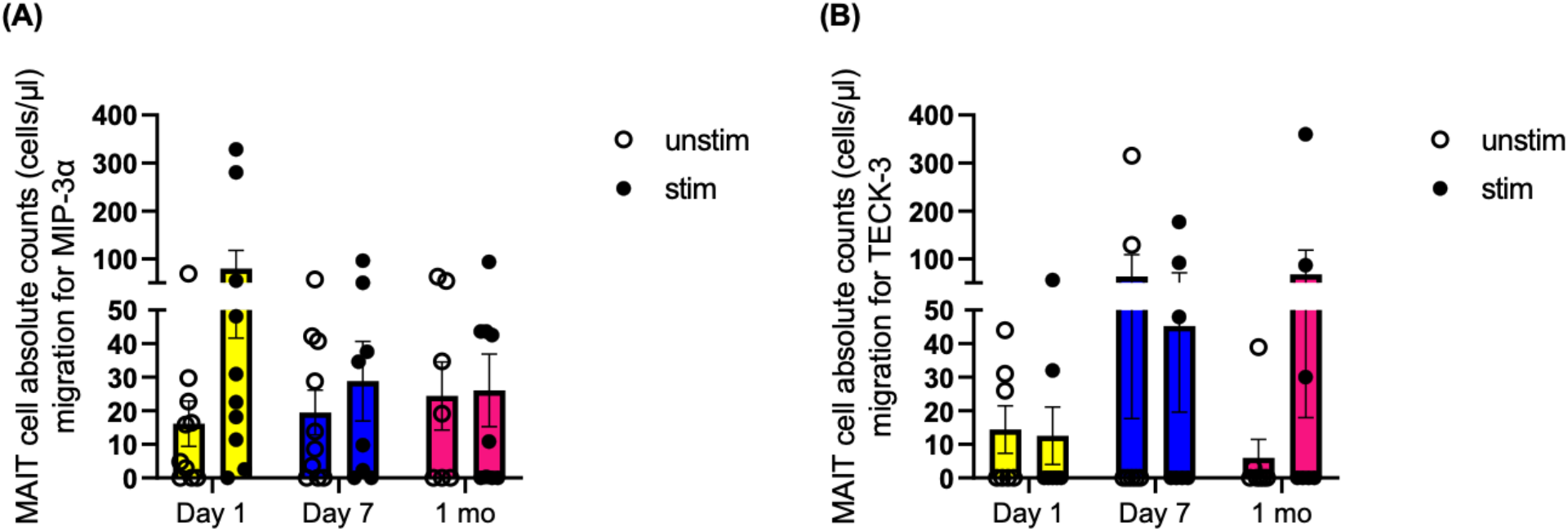
MAIT cell migration towards chemokines MIP-3α and TECK-3. PBMCs from vaccine recipients were seeded on upper chamber of 3-μm pore transwell with indicated chemokines at 150 ng/mL in the bottom well and were allowed to migrate for 4 hours at 37°C. Absolute numbers of MAIT cells migrated towards **(A)** MIP-3α and **(B)** TECK-3 chemokines were quantified using flow cytometry. Mean and SEM of two experiments are shown. \**P* < .05, \*\**P* < .01, \*\*\**P* < .001 in Wilcoxon signed-rank test (paired samples).

